# Modeling the Effects of Non-Pharmaceutical Interventions on COVID-19 Spread in Kenya

**DOI:** 10.1101/2020.05.14.20102087

**Authors:** Duncan K. Gathungu, Viona N. Ojiambo, Mark E. M. Kimathi, Samuel M. Mwalili

**Affiliations:** Jomo Kenyatta University of Agriculture and Technology; Machakos Univesity

**Keywords:** Basic reproduction number, COVID-19, SEIR, NPIs

## Abstract

May 14, 2020

Mathematical modeling of non-pharmaceutical interventions (NPIs) of COVID-19 in Kenya is presented. An SEIR compartment model is considered with additional compartments of hospitalized population whose condition is severe or critical and also the fatalities compartment. The basic reproduction number (*R*_0_) is computed by next generation matrix approach and later expressed as a time-dependent function so as to incorporate the NPIs into the model. The resulting system of ordinary differential equations (ODEs) are solved using fourth-order and fifth-order Runge-Kutta methods. Different intervention scenarios are considered and results show that, implementation of closure of education insitutions, curfew and partial lockdown yield predicted delayed peaks of the overall infections, severe cases and fatalities and subsequently containement of the pandemic in the country.

## 1 Introduction

On 7th January 2020, the World Health Organization (WHO) reported novel severe acute respiratory syndrome coronavirus (SARS-CoV-2) virus causative of the COVID-19, [4]. The WHO report, details the chrolonology of how Wuhan in Hubei Province in China became a global epicenter of COVID-19 with the epidemiological link to the Huanan Seafood wet markets where there was sale of live animals. Over the period, COVID-19 has infected close to 3 million people worldwide with close to 200,000 fatalities reported, necessitating the WHO to declare it a pandemic. With no viable vaccine at the moment, COVID-19 has initiated efforts focussed on containement and strategies aimed at reducing infections and fatalities. On the global platform, there has been a furor of activity in determining the best strategy in containment of the COVID-19 pandemic. A host of countries have insituted interventions not limited to social distancing, curfews and total lockdowns. In [3], a model taking into consideration Bats-Hosts-Reservoir-People transmission network was implemented, where reproduction numbers (*R_O_*) were computed to determine the transmissibility of the virus between people and reservoirs. The value of *R_O_* of reservoir to people was found to be 2.30 while people to people was 3.5. In [11], they delved into mathematical models of the COVID-19 and with a reproduction number of 2, they considered an SEI model to model the effects of local social gatherings. With a 14-day infectious period, the model showed that with 18 hours exposure, the attendees of the event had a protection efficacy of 70%. In [8], a conceptual SEIR model to gauge the effects of individual reaction and goverment reactions was investigated. This was in line with Chinese government implementation of social distancing measures in areas considered as epicenters. The model incorporated zoonotic introductions and emigrations and showed that close relations with reported cases, even though asymptomatic transmission could not be established. In Ontario, Canada, [13] implemented a mathematical model for transmission and mitigation strategies. They considered measures taken in the population which included quarantine, isolation of infectious cases and hospitalization with intensive case unit (ICU) cases. It was reported that in absence of substantial physical social distancing and enhanced case detection and isolation, the ICU resources will be quickly overwhelmed. In [1], considered a model for prediction of new cases ad made a comparison study of United States of America, India and Italy. Different scenarions under physical distancing measures such as lockdown were considered and predictions were made using the available data. It was reported that, without mitigation of physical interaction of the populations, the peaks of infections will be realised much earlier in all the three countries.

Of late the modeling efforts have advanced to cater for age-structured modelling to determine the transmission across different age stratas. In [12], they use an age-structured SIR model with social contact matrices from surveys and Bayesian imputation to gauge the spread of COVID-19 epidemic in India, where they use a generalization of the time dependent *R_O_* case study data, age distribution and social contact structure. Their predictions implore on the duration of social distancing mitigation such lockdowns and give scope of when periodic ease of the lockdowns can be implemented. Further age-structured morbidity and mortality is reduced on implementation of these measures. [5] investigated a more complex variant of the SEIR referred to as θ−SEIHRD model to take into account the undetected infections and based their study in China. Their study showed that mitigation measures of limiting physical contact decrease the basic reproduction number and subsequently a reduction in infections. Further brisk detection is needed in order to reduce the number of infections. In [6], results of social distancing strategies for curbing the COVID-19 epidemic are presented. They simulate the reduction of reproduction numbers as a result of social distancing measures. In [7], an important parameter referred to as the identification parameter is included in the simulations to predict the peaks of the epidemic in Japan. This parameter is the ratio between the positive cases to the number of tests performed. Further they used available date and the least-square based procedure with Poisson noise to estimate the infection rate as a function of the identification rate and they consider an SEIR compartmental model. They report that, interventions have a positive effect in delaying the peak of the epidemic and they propose longer interventions to contain the epidemic.

The results of the researches outlined, provide techniques knowledge on implemeting a mathematical model for the epidemic in Kenya. Recently, [2] outline the forecast of the scale of the COVID-19 epidemic in Kenya. In the study they investigate the dissimilarities between China and Kenya interms of the demographics and geography. They forecasted the potential incidence rate and magnitude of the epidemic using observed growth rate and age-distribution of confirmed cases in China. They reported that the number of the infected cases is likely to be high and that isolation of the asymptomatic infectious population will not be enough measure. Further they proposed exceptional social distancing to ’flatten the curve’ and caution on the expected rebound of infections when the restrictions are lifted. In [9], an SEIR model was implemented taking into consideration the population’s interaction with the enviroment. This was achieved by considering the enviroment as the host of SARS-CoV-2 virus. In the absence of social distancing, transmission and spread of COVID-19 was rife.

As at 7th May 2020, Kenya had performed 28,002 tests, with 607 confirmed positive of COVID-19, 197 recoveries and 29 fatalities, [10]. The country’s peak is not yet predicted and in this paper, we investigate a variant of the SEIR model to predict the peaks of the infections, severity of the ill and fatalities with and without the non-pharmaceutical measures.

## 2 Model formulation

To model the mitigation efforts to curb the transmission and spread of COVID-19, we use the compartmental Susceptibles-Exposed-Infected-Recovered model. We consider human-human transmission and divide the human population *N* at a time *t* into eight compartments. The susceptible population is denoted by *S*(*t*), the exposed population by *E*(*t*), the asymptomatic infectious population by *A*(*t*), the mild symptomatic population by *M*(*t*), the population who are severe and hospitalised by *H*(*t*), the critically ill population in the intensive care unit (ICU) by *C*(*t*), the recovered population by *R*(*t*) and the fatalities by *D*(*t*). Hence the total human population is given as

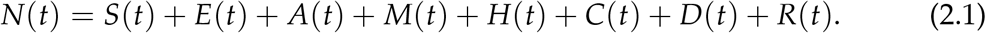

*S*(*t*) is the human population in Kenya that maybe infected with (SARS-CoV-2) virus. The susceptibles move to *E*(*t*) after incubation period to the onset of the disease. The exposed individuals move to either either asymptomatic *A*(*t*) or the mild symptomatic *M*(*t*). Assuming the population in the asymptomatic have ’immunocompetency’, they move to *R*(*t*). The population from *M*(*t*) can move either to *R*(*t*) or if their conditions deteriorate warranting hospitalization they move to *H*(*t*) and are considered severe. The severe population on assessment can move to critical compartment *C*(*t*) and if their condition improve and become less critical, they are referred back to the general ward as severe cases. Further on improved conditions they recover and move to *R*(*t*). The population in *C*(*t*) who succumb move to the *D*(*t*).

On 13th March 2020, after the first confirmed case, the Government of Kenya (GOK) instituted measures towards addressing the spread and transmission of COVID-19. The measures and interventions initially include a dusk-dawn curfew and massive campaign and sensitization measures of social and physical distancing. On 6th April 2020, the GOK extend and upscaled these measures with partial lockdowns in towns considered as hotspots in Kenya. These measures constituted enabled formulation of assumptions for development of the mathematical model. We assume the following;-

1. The disease is transmitted through human-human transmissions. There are no cross-infection occurring from pathogens in the environment nor human-animal transmissions.
2. Susceptible (S) individuals are exposed/infected through contact with infectious individuals. Each infectious individual causes an average *R*_0_ secondary infections.
3. After an average incubation period of 5.1 days, exposed (E) individuals either become asymptomatic (A) or exhibit mild infections (M), but not all infected person exhibit symptoms.
4. The virus-infected person is not infectious during the incubation period.
5. Individuals with mild infection either recover (R) or worsens to a severe case (H).
6. Individuals with severe infection either recover (R) or worsens to a critical case (C).
7. Critically ill individuals either return to regular hospital or die.
8. Demographics such as birth, death rates and immigration were not considered.
9. Only a fraction of infective individuals can be identified by diagnosis.
10. An infected individual acquires immunity upon recovery.

The flowchart below illustrates the compartmental SEIR model under consideration.

The variant of the SEIR compartmental model illustrated by the Figure 1, culminates to an eight-dimensional dynamical system of ordinary differential equations(ODEs) given by,

**Figure 1:**
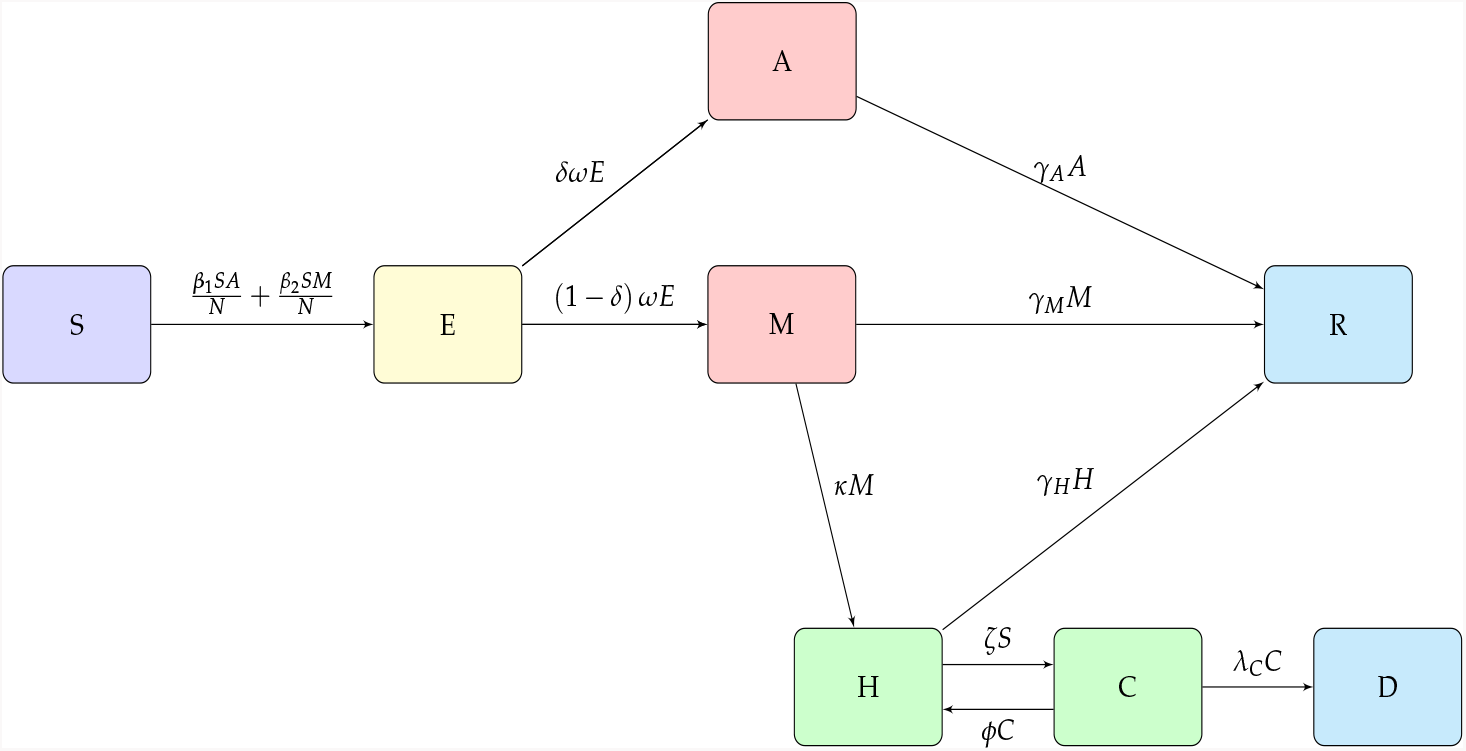
SEIR model flowchart

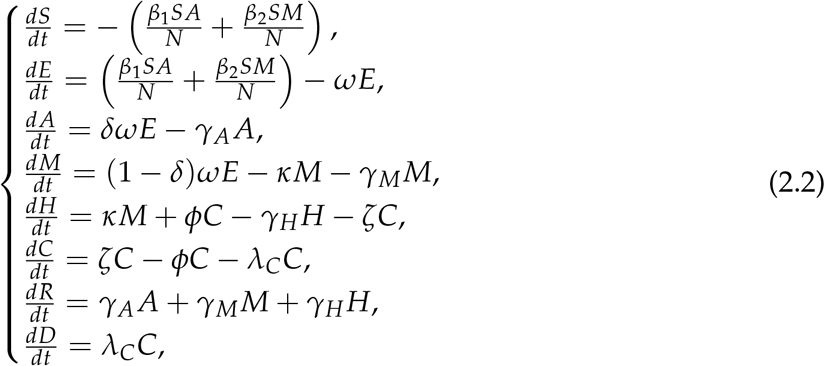

with the initial conditions:

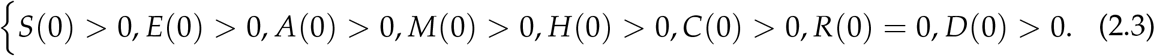

The parameters used in the model are given in Table 1.

**Table 1:**
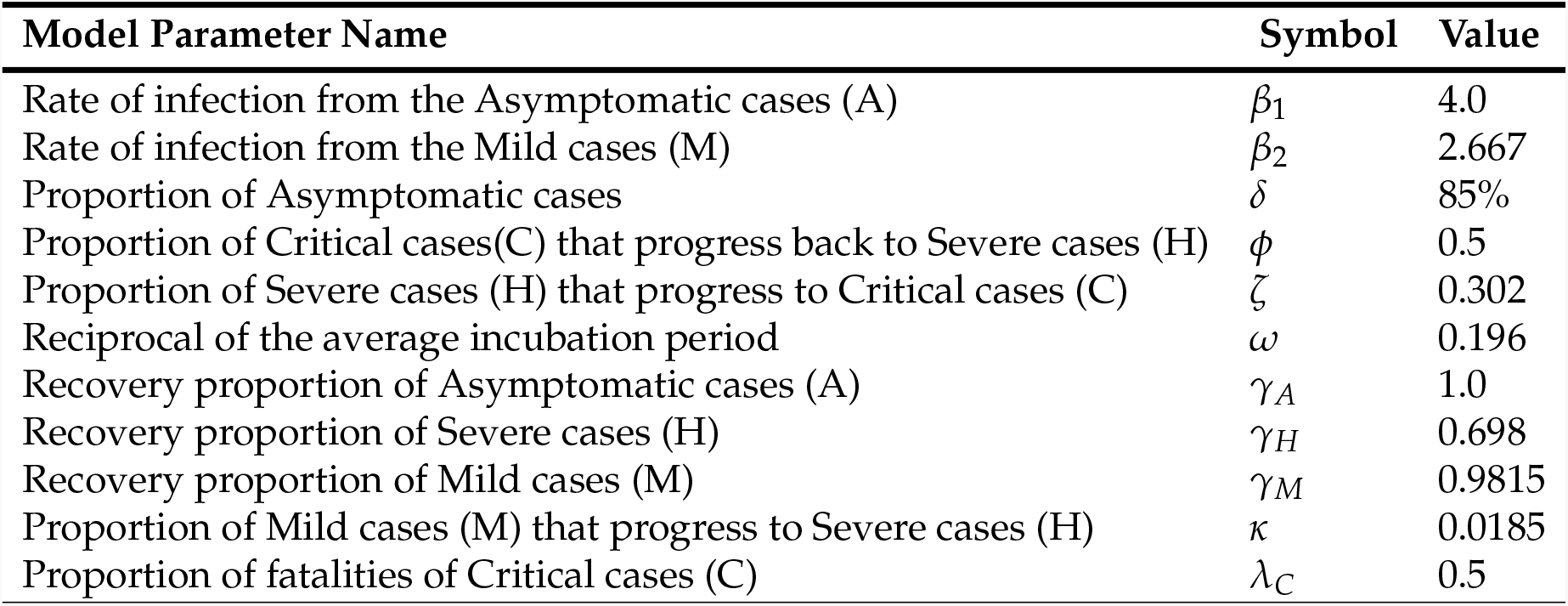
Parameters of the SEIR model of COVID-19

| Model Parameter Name | Symbol | Value |
| --- | --- | --- |
| Rate of infection from the Asymptomatic cases (A) | $\beta_1$ | 4.0 |
| Rate of infection from the Mild cases (M) | $\beta_2$ | 2.667 |
| Proportion of Asymptomatic cases | $\delta$ | 85% |
| Proportion of Critical cases(C) that progress back to Severe cases (H) | $\phi$ | 0.5 |
| Proportion of Severe cases (H) that progress to Critical cases (C) | $\zeta$ | 0.302 |
| Reciprocal of the average incubation period | $\omega$ | 0.196 |
| Recovery proportion of Asymptomatic cases (A) | $\gamma_A$ | 1.0 |
| Recovery proportion of Severe cases (H) | $\gamma_H$ | 0.698 |
| Recovery proportion of Mild cases (M) | $\gamma_M$ | 0.9815 |
| Proportion of Mild cases (M) that progress to Severe cases (H) | $\kappa$ | 0.0185 |
| Proportion of fatalities of Critical cases (C) | $\lambda_C$ | 0.5 |

The terms 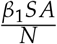 and 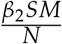 describe the rate at which the susceptible population are infected by the asymptomatic population and the mild symptomatic respectively.

In order to gauge the extent of the spread of COVID-19, experts have recommended extensive testing in the populations but due to economic constaints in the country and globally this testing has been below par. Therefore, many countries have resulted to introducing a wide range of NPIs to atleast slow down the epidemic spread as they gather resources for mass testing and isolation of confirmed cases. In order to simulate and predict infection cases from the model, we use the framework developed by [7]. In this case it is assumed only a fraction of the infected individuals are identified by the diagnosis. This fraction is given by *p* and it is referred to as the identification parameter which is 0 *< p* 1. For this study *p* is calculated as the ratio of the confirmed cases to the tests performed. Its usage in this study is as follows: suppose *S* + *E* + *A* + *M* +*H* + *C* + *R* + *D* = 1, and one person is infected in Kenya of population 4.76 *×* 10^7^ at time *t* = 0. Then the infectious individuals *A*(0) + *M*(0)are given by *In f* (0) = *p*(*A*(0)+*M*(0))=1, hence 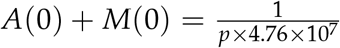. At any time *t* the number of infectious individuals is given by *In f* (*t*) = *p* (*A*(*t*) + *M*(*t*) *×* 4.76 *×* 10^7^ = Further at *t* = 0, if we assume *E*(0) = *H*(0) = *C*(0) = *R*(0) = *D*(0) = 0, then

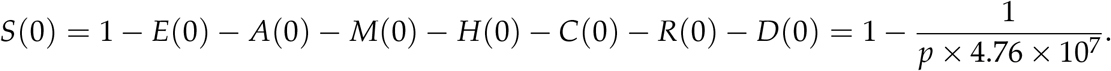

## Existence of a disease free equilibrium(DFE)

To determine the DFE of the COVID-19 in Kenya we solve the system of equations (2.2) after equating the right hand side of the system to zeros. In this case *A* = 0, *M* = 0, *H* = 0 and subsequently *C* = 0, *R* = 0 and *D* = 0, which yields a DFE point as

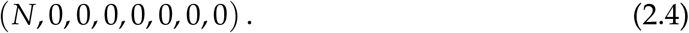

We compute the basic reproduction number *R_O_* at DFE using the next generation approach. Let *x* = (*E, A, M*)^*T*^ and 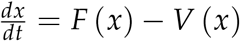, where;

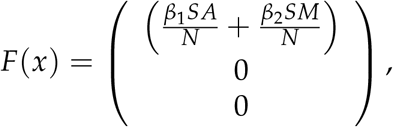

and

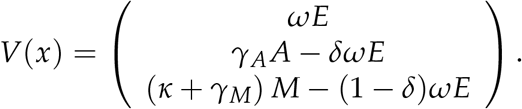

Obtaining the derivatives of *F*(*x*) and *V*(*x*) at DFE point yields **F** and **V** matrices as follows

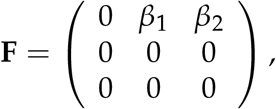

and

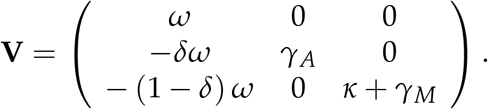

The *R*_0_ is the spectral radius of the product of **FV**^*−*1^;

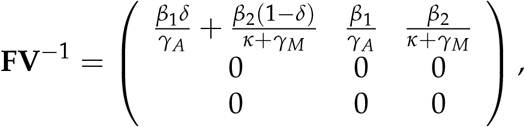

where *R*_0_ is given as;

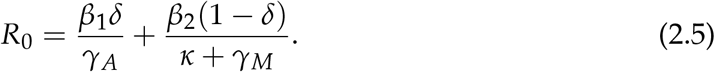

This *R*_0_ consists of two terms showing transmission by the asymptomatic population and the mild symptomatic population.

## Existence of Endemic Equilibrium Point(EE)

The endemic equilibrium point of the model (2.2) is determined as positive steady state in which the COVID-19 is said to persist in a given population.

**Theorem 1**. *The COVID-19 model has a unique endemic equuilibrium point when R*_0_ *>* 1 *otherwise the endemic equilibrium doesn’t exist*.

**Proof**. Suppose that (*S^*^, E^*^, A^*^, M^*^, H^*^, C^*^, R^*^, D^*^*) is a non-trivial equilibrium point of (2.2). The equilibrium (*S^*^, E^*^, A^*^, M^*^, H^*^, C^*^, R^*^, D^*^*) is determined by solving the equations;

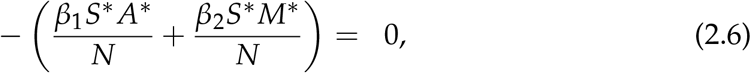

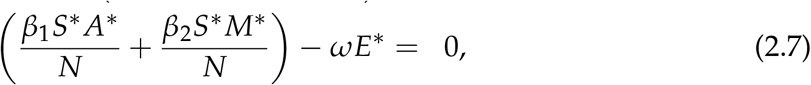

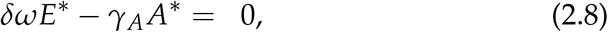

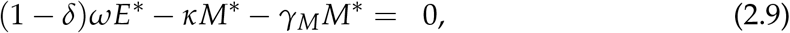

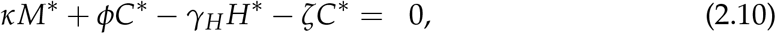

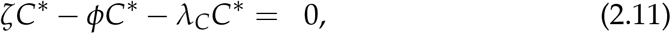

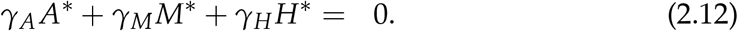

From (2.8) and (2.9) we get;

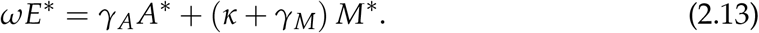

From (2.11) we get

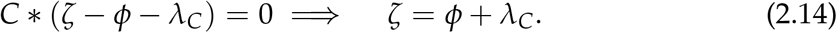

From (2.12), we get;

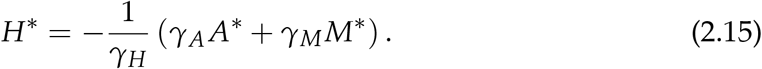

From (2.10) we get

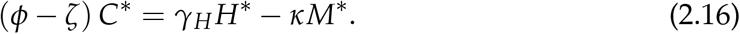

Using (2.15) in (2.16) yields;

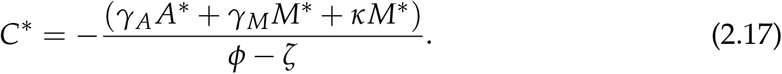

From (2.7) we get;

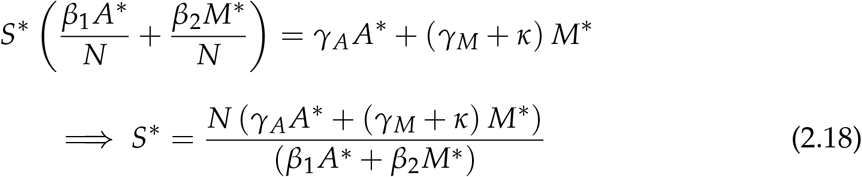

Both *A*\* and *M*\* should satisfy

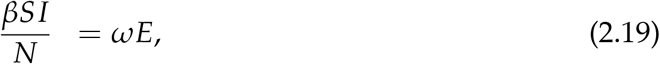

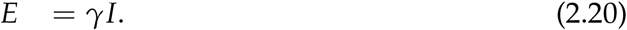

Using (2.20) in (2.19) yields 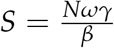

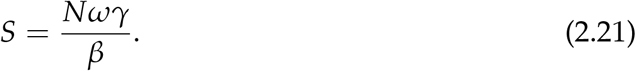

Using (2.20) in (2.13) yields;

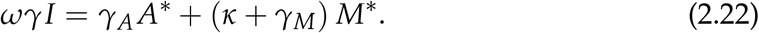

Using (2.21) in (2.18) yields;

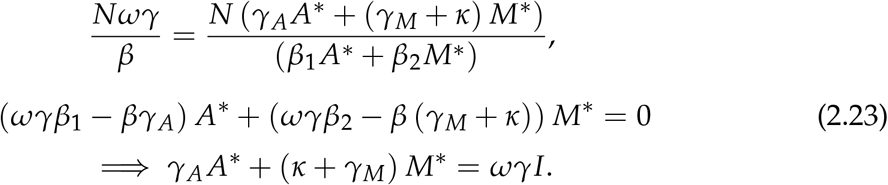

In an endemic scenarion *I ≈ αN*, where *α* is the attack rate, since almost everyone gets infected. Therefore solving the following simultaneously yields *A*^*^ and *M*^*^.

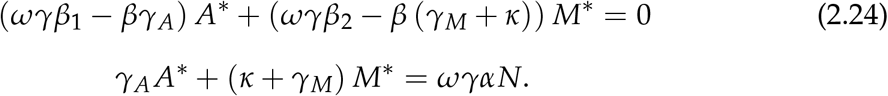

hence

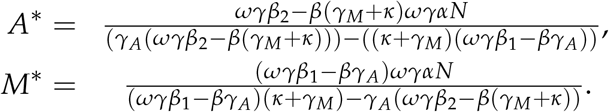

## Prediction of infection cases

In the quest to determine the level of preparedness to curb spread of COVID-19, it is important to predict future numbers of daily infection cases and cumulative infections. This provides a basis for policy makers to know when the available healthcare infrastructure is likely to be overwhelmed. In this section, we used a Metropolis-Hastings-Markov-Chain-Monte Carlo (MCMC) algorithm for non-linear Gaussian functions to estimate the daily and cumulative cases from the Kenya’s data of daily confirmed cases. Using 20,000 iterations of MCMC procedure we are able to predict the daily infection cases and the cumulative infections for a period of 90 days.

Using the available daily data of the COVID-19 cases, [10], we are able to use these procedures to predict the daily infection cases as seen in Figure 2a and when the country is likely to have reported the first 1000 infection cases from Figure 2b.

**Figure 2:**
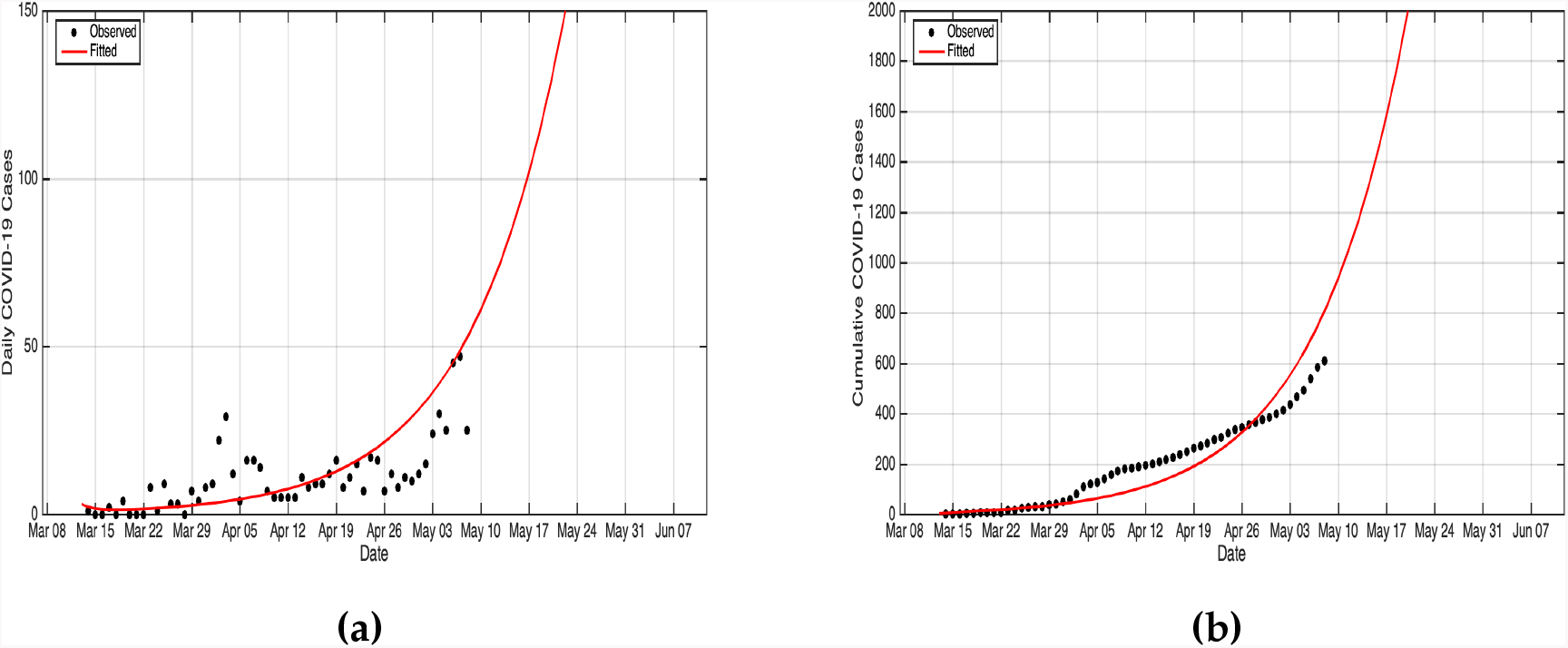
(a) Daily cases prediction over a 90 days period (b) Prediction of the cumulative cases over a 90 days period.

## 3 Simulation and Results

In this section, we report on the results of simulations of COVID-19 model. The model’s equation 2.2 that are solved using a fourth and fifth order Runge-Kutta method, which is implemented in MATLAB. We consider *N* = 4.76 × 10^7^ people which is Kenya’s total population as per the 2019 census. The simulations are done using the parameters listed in Table (1). The human-human contact reduction measures we consider are school closedown, dusk to dawn curfew and partial lockdowns in towns and cities perceived as COVID-19 hotspots. The essence of contact reduction is to mitigate the interaction between the susceptible and infectious populations. Based on studies of COVID-19 spread in China, Italy, and Spain, we suppose that if no mitigation measures are in place then an infectious individual would infect three secondary cases, in his/her interaction sphere. Thus we consider the *R*_0_ as 3, for the unmitigated scenario.The first case of the novel corona virus was confirmed in Kenya on 13th March, 2020. On 16th March 2020, schools were closed, which was the first NPI that was implemented by the Kenyan government. Then 14 days into the school closedown, curfew measures were implemented. Thereafter 24 days into school closedown, partial lock-down was implemented. In our simulations, we implement the school closure for 210 days, curfew for 196 days, and partial lockdown for 186 days. This is done via a time-dependent Ro, which was taken as a cosine function, see [6]. We presume that school closure yields a 20% reduction in contact while adding a curfew on to the school closure results to a 40% reduction, and introduction of the partial lockdown on to the other two NPIs yields a 60% reduction in contacts as shown in Figure 3.

**Figure 3:**
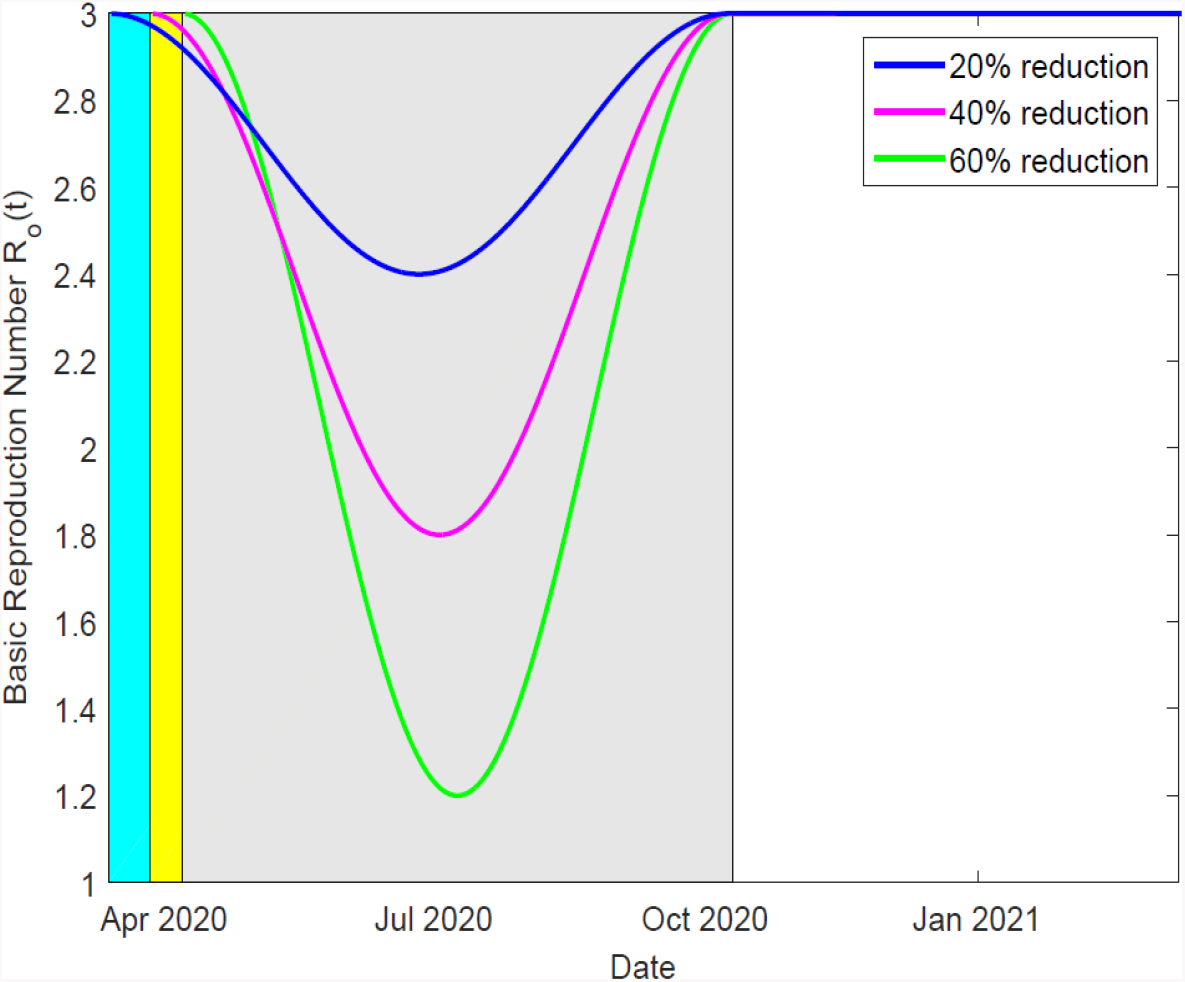
Depiction of the effects of school closedown, curfew and partial lockdown on the time-dependent basic reproduction number. The duration of school closedown, of 210 days, is indicated by the cyan color. However, it is overlapped by the duration of curfew implementation, shown here in yellow. The yellow region is in turn overlapped by the gray region which represents the duration of partial lockdown implementation.

A reduction in *R*_0_ implies that fewer individuals are being infected and hence the disease spread is slowed down.

In Figures 4a and 4b, we report on the simulation of infections and the cumulative infections under different scenarios. These different scenarios are when there is no any mitigation, when the schools are closed, when schools are closed and dusk-dawn curfew is instituted and when schools are closed, curfew in place and partial lockdown is in place.

**Figure 4:**
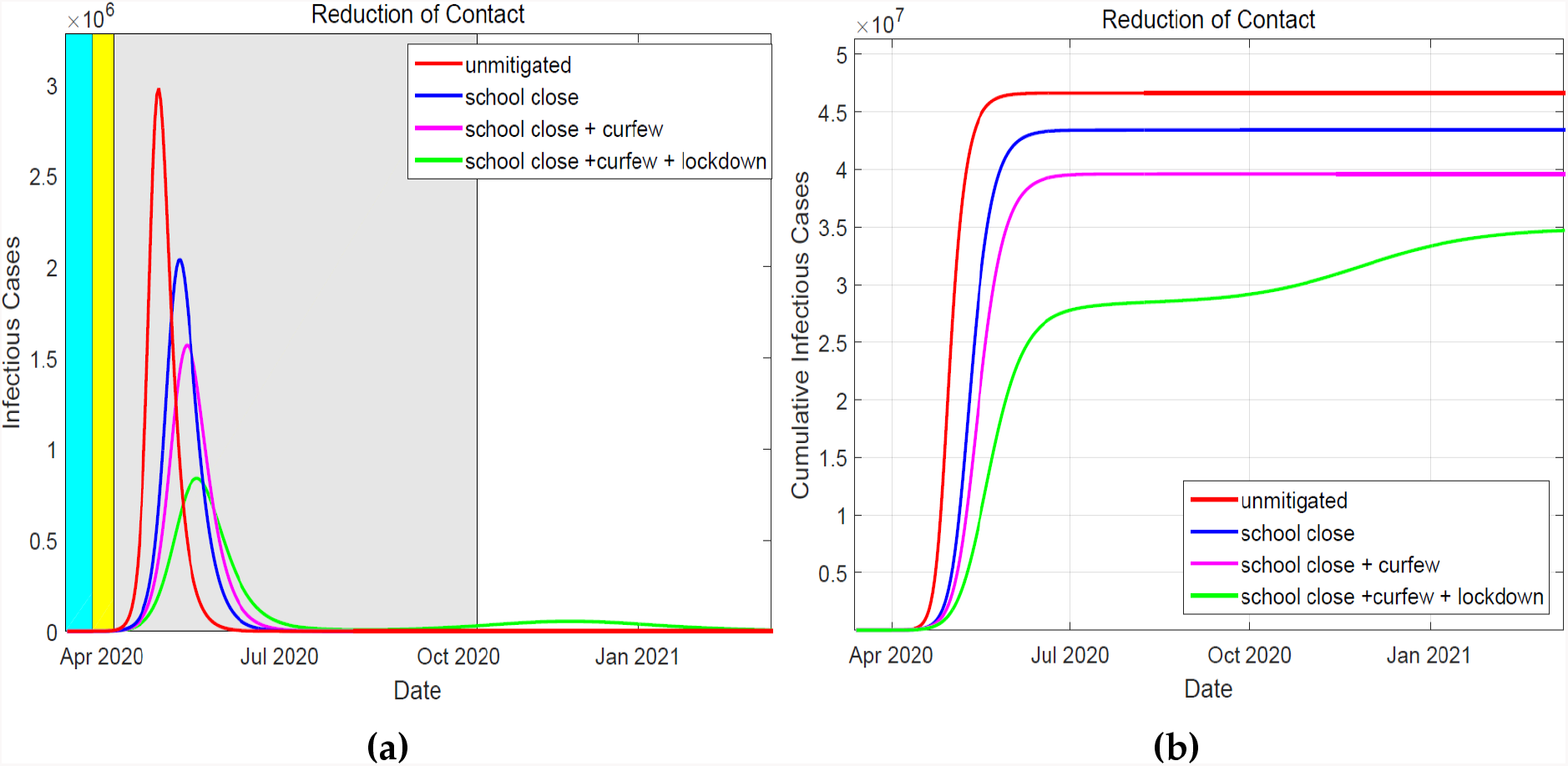
The simulated daily cases of infected individuals is shown in a), whereby the duration of NPIs implementation is also indicated by the colored regions. The cumulative cases of the infected individuals is depicted in b). The NPIs are shown to effectively mitigate the spread of the epidemic in the population. A combination of the three measures is the most effective strategy in that even though a new wave of infection emerges, it flattens out with a much lower peak.

From the figures we deduce that, the NPIs instituted aid in containment of the epidemic spread, resulting to fewer people being infected as compared to the unmitigated scenario. In particular, we see a much higher peak of infections in the unmitigated scenario, which arrives much faster due to the high rate of infections when *R*_0_ = 3.0. For the first two NPIs the infections peaks are lower and come at a later date. When all the three NPIs are combined, resulting to a *R*_0_ of 1.2, the infections peak at a much lower value and the epidemic begins to subside. However, this leaves a large population in the susceptible compartment such that when the value of *R*_0_ begins to rise, towards the end of the NPI duration, a rebound of infections occurs. Due to the effectiveness of the combined NPIs the new peak is much lower and the epidemic is eradicated. The measures results to a reduction in the cumulative number of infections, see Figure 4b. Combining all the three NPIs leads to a significant delay of the epidemic spread, a time the health system can utilize to put mechanisms and facilities to respond to the epidemic appropriately.

In Figures 5a and 5b, we report on the number of severe cases of COVID-19 who are hospitalized. Reduction of the severe cases is important as this represents proportion of the population who are directly utilizing the healthcare facilities at a given time. Due to limited hospital infrastructure and resources, delay of this peak is vital for the health systems to prepare for a guaranteed influx of patients in need of medical attention. Furthermore, any reduction of the hospitalized results in significant reduction of severe cases transfer to critical cases that require ICU services.

**Figure 5:**
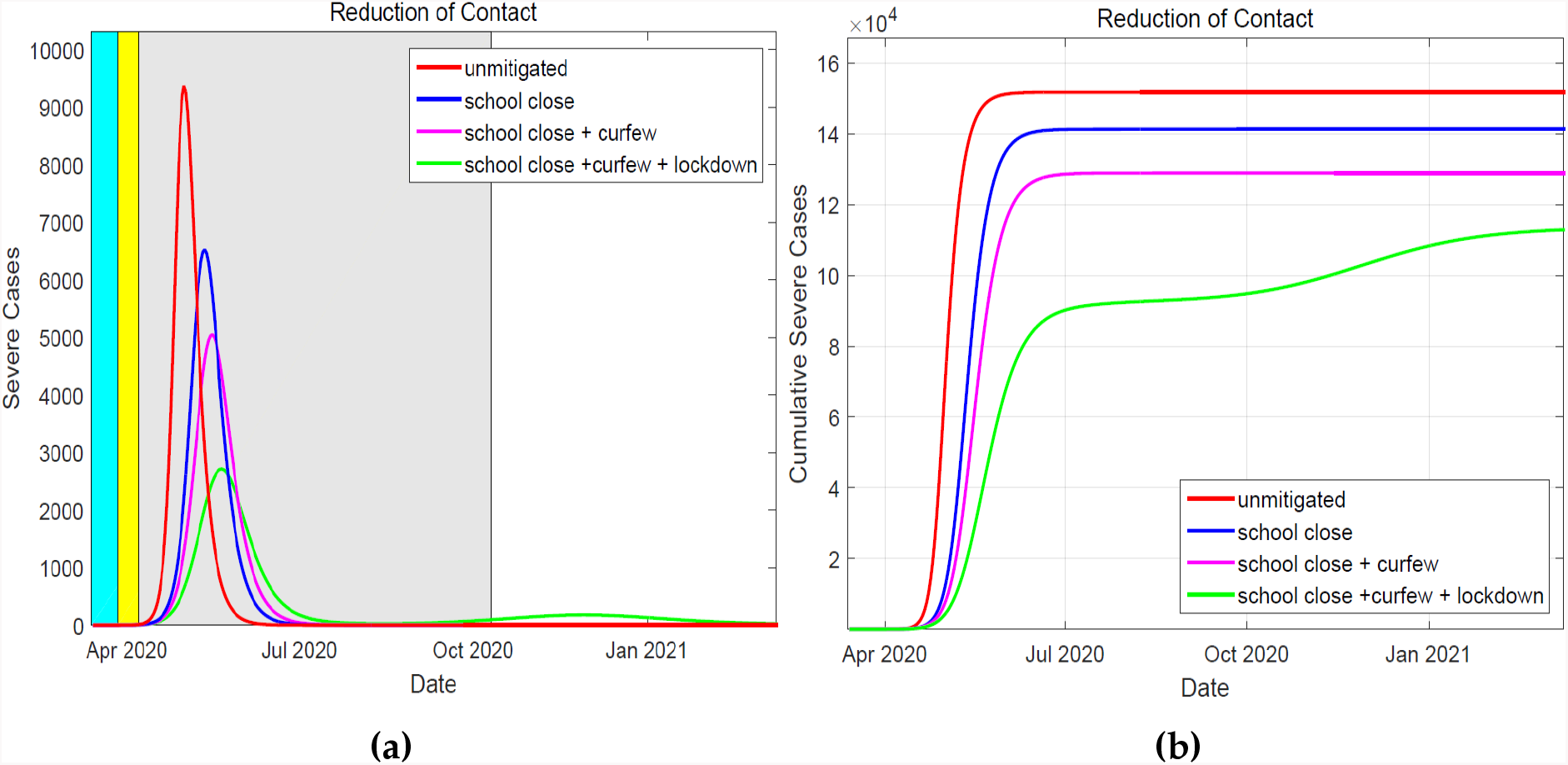
Over the duration of the implementation of the NPIs, the daily severe cases are shown in (a) where the duration of these interventions is indicated by the colored region. In (b), the cumulative severe cases for the three mitigation measures are shown. It shows that, the implementation of the three measures lead to a reduced number of severe cases.

From the Figures 5a and 5b, we deduce that strigent implementation of the NPIs is key in ensuring the available hospital infrastructure is not overwhelmed during the epidemic. The national goverment and county goverments under the Health ministry directive have boosted the bed capacity at all hospital levels, including establishing isolation centers. If schools are closed, curfew implemented and lockdown instituted, the simulation shows that the hospitalized population may not overwhelm the established healthcare infrastructure capacity.

In Figures 6a and 6b, we present the results of simulations of fatalities under different intervention scenarios.

**Figure 6:**
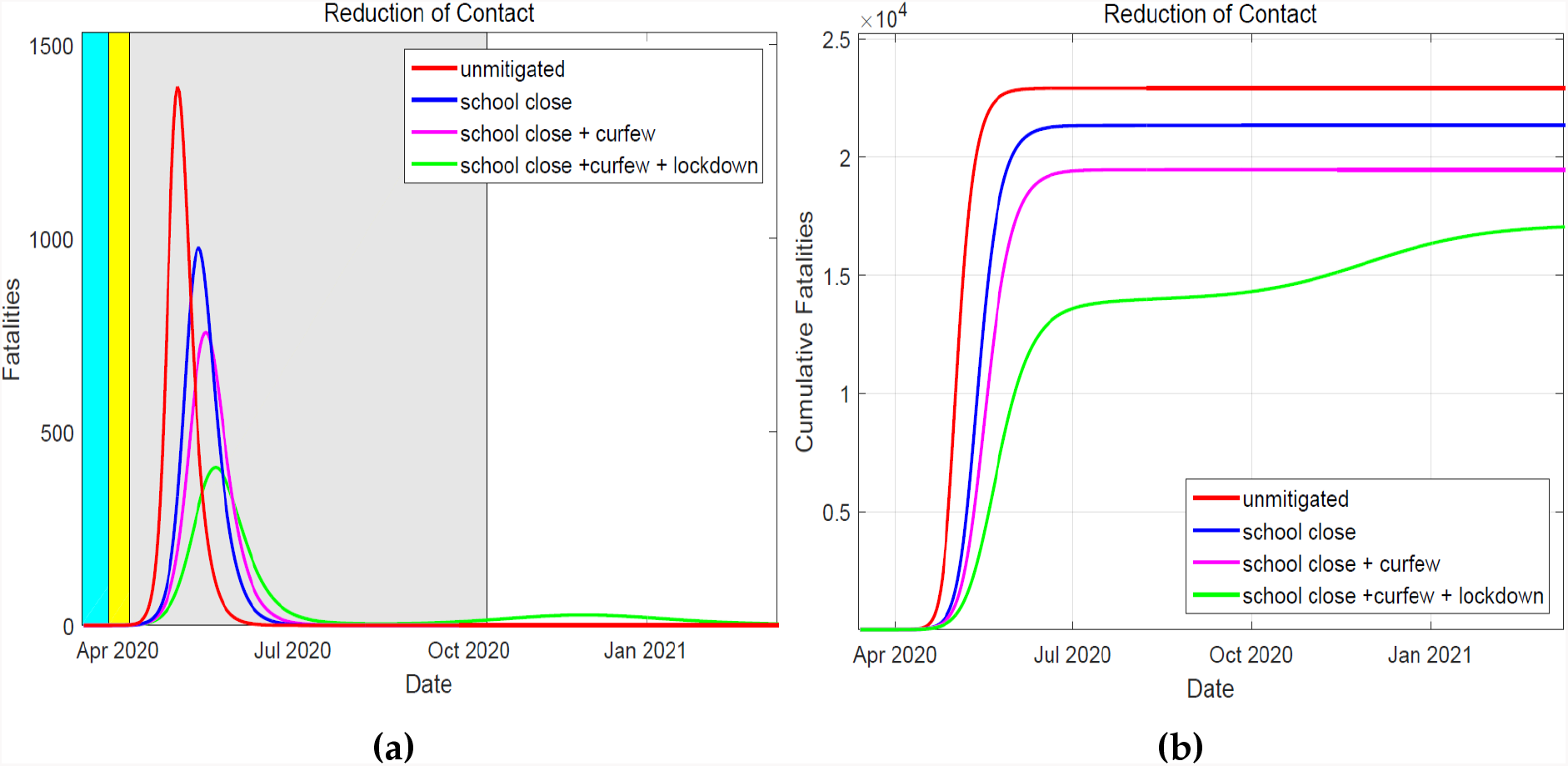
The simulated daily number of fatalities is shown in (a), where the duration of the NPIs is indicated by the colored region. The cumulative number of the fatalities is presented in (b). In the event all three mititgation measures are in place, the cumulative cases ‘plateau’ at a much lower peak.

Obviously, fatalities from COVID-19 will have the highest peak if NPIs are not instituted. From the Figure 6a, delayed peaks are realised if the NPIs are in place and the best scenario is if the considered measures are combined, for the considered duration of implementation. A rebound of infections also results to a resurgence of fatalities due to the severity of the disease, especially among the older population.

## Conclusion

Since COVID-19 vaccine development will take significantly longer period of time before it becomes globally available, it is imperative on all medical sectors in the country to adhere to mitigation measures and intervention policies set by the government. The implementation of school closure, dusk to dawn curfew and partial lockdown are the mitigation effective in curbing the spread of COVID-19 that are effective and lead to the flattening of the curve within the duration of implementation. Relaxation of the mitigation measures on or before September 2020 will likely lead to a resurgence and the country may experience a new wave of infections. Devoid of the aftershocks of these interventions, the country will have no infections as at 31st December 2020.

## Data Availability

The data used to support findings of this study is available from the corresponding author upon request.

https://www.health.go.ke/

